# An Agentic, No Code Artificial Intelligence Workflow for Developing and Externally Validating a Thyroid Nodule Ultrasound Malignancy Classifier

**DOI:** 10.64898/2026.06.23.26356395

**Authors:** Johnson Thomas, Nikita Pozdeyev

**Author notes:** Corresponding author: Johnson Thomas, MD, FACE — Department of Endocrinology, Mercy Hospital, Springfield, Missouri, USA.

## Abstract

Convolutional neural networks (CNNs) can classify thyroid nodules on ultrasound, yet published models are seldom available for independent testing, require machine-learning expertise to develop and deploy, and are validated mostly on papillary thyroid carcinoma.

**Objective:** To test whether an autonomous (“agentic”), no-code artificial intelligence (AI) agent can develop a calibrated thyroid-nodule malignancy classifier, and to validate it internally and on an external cohort spanning multiple cancer histologies.

**Methods:** This is a retrospective, computational diagnostic study with prespecified endpoints. A no-code agent (Hugging Face ML-Intern) autonomously reviewed data, selected and trained the model and calibrated probabilities, using the open-source TN5000 dataset (3500 training, 500 validation, and 1000 test images). The trained ResNet-18 model was externally validated on 232 nodules from the University of Colorado, including follicular, medullary, oncocytic, and follicular-variant of papillary carcinomas.

**Results:** On the internal test set, an agentic AI model achieved AUROC 0.94 (95% CI, 0.920–0.953), sensitivity 0.90, and specificity 0.80. On external validation, agentic AI model achieved an AUROC of 0.90 (95% CI, 0.850–0.936), sensitivity of 0.92, and specificity of 0.68, negative predictive value of 0.96, and positive predictive value of 0.52, exceeding the performance of a previously published classifier on the same cohort (AUROC of 0.83).

**Conclusions:** An agentic, no-code AI workflow produced a calibrated, externally validated thyroid nodule classifier, supporting accessible, reproducible, and independently testable medical AI development. Prospective validation and local recalibration are required before clinical use.

## Introduction

Artificial intelligence (AI) methods for thyroid nodule ultrasound analysis have expanded rapidly, with hundreds of publications describing models for nodule detection, segmentation, sonographic characterization, risk stratification, and malignancy prediction^1–3^. However, a persistent translational problem, i.e., most thyroid nodule AI tools remain difficult for clinician researchers to independently test, reproduce, compare, or deploy, remain unsolved. This gap is clinically important because thyroid nodules are common, detected on high-resolution ultrasound in up to 68% of randomly selected adults, yet most are benign. Current risk-stratification systems, including the American College of Radiology Thyroid Imaging Reporting and Data System (ACR TI-RADS) and the American Thyroid Association (ATA) risk patterns, rely on subjective interpretation of sonographic features.^4,5^ Interobserver variability in ultrasound feature assessment can alter thyroid nodule risk categorization and fine-needle aspiration (FNA) recommendations. Improving the consistency and specificity of ultrasound-based triage may reduce potentially avoidable FNAs of thyroid nodules that are unlikely to be malignant.^6,7^

Deep convolutional neural networks (CNNs) applied to thyroid ultrasound have shown diagnostic performance that can approach, and in some settings match, that of expert radiologists for distinguishing benign from malignant nodules^8,9^. Several commercial and FDA-cleared AI-enabled tools for thyroid ultrasound have also become available^10^. Nevertheless, three practical barriers limit translation. First, many published models are not made publicly available for independent testing or head-to-head benchmarking, preventing clinicians from evaluating performance on their own institutional data. Second, even when code is released, model deployment typically requires configuring a machine-learning environment, preprocessing images, managing dependencies, and writing inference scripts—tasks that are beyond the technical reach of many clinician investigators. Third, most thyroid ultrasound AI models have been developed and evaluated primarily in datasets dominated by papillary thyroid carcinoma (PTC), with limited assessment in follicular-pattern and non-PTC malignancies, including follicular thyroid carcinoma (FTC), follicular variant PTC (FV-PTC), oncocytic carcinoma (HCC), and medullary thyroid carcinoma (MTC).^3,11^ These cancers may lack the classic suspicious sonographic features on which both clinical risk-stratification systems and image-based machine-learning classifiers often depend. As a result, it is difficult for institutions with ultrasound datasets, which may be acquired using varied scanners, acquisition protocols, and consist of various thyroid cancer histologies, to determine whether published AI models generalize to their local populations and are usable clinically.

Agentic AI, large language model (LLM)-based systems that can autonomously coordinate multistep computational workflows may lower this barrier by allowing nonprogrammers to perform model development tasks that traditionally require machine-learning expertise^12^. In principle, an agentic AI system could inspect a dataset, select and configure a model, train and calibrate the classifier, evaluate performance, and generate deployable inference code with minimal manual programming. Such systems could enable clinician-researchers who possess thyroid ultrasound data but lack coding expertise to develop and test institution-specific classifiers. To our knowledge, autonomous no-code agentic development of a thyroid ultrasound malignancy classifier has not been rigorously evaluated.

We used a publicly accessible no-code agentic AI platform, Hugging Face ML-Intern^13^, to autonomously develop a calibrated ResNet-18 thyroid ultrasound malignancy classifier using the open-source TN5000 dataset^14^. We then: (i) evaluated the locked model on a prespecified internal test set; (ii) externally validated it on an independent University of Colorado cohort that includes non-papillary thyroid cancer histologies^15^; (iii) benchmarked model’s performance against a previously published classifier evaluated on the same external source; and (iv) developed a freely testable, browser-based on-device deployment to support transparent model evaluation by clinician researchers.

## Materials and Methods

### Study design and ethics

This was a retrospective, computational diagnostic accuracy study using only publicly available, fully de-identified data and publicly accessible cloud computing. Patients were not recruited, contacted, or consented, and protected health information was not accessed. The study was determined to be exempt by the Mercy Institutional Review Board. The external University of Colorado data originated from a prior study and is publicly accessible. Thyroid ultrasound imaging data collection at the University of Colorado was approved by the Colorado Multiple Institutional Review Board under protocol #20-2315.^15^The analysis plan, including the model selection, metrics and calibration, was specified before the test set was examined; the verbatim natural-language prompt that defined the workflow is provided in Supplementary Material.

### Agentic AI platform and reproducible workflow

Model development was performed entirely through the ML-Intern.^13^From a single natural-language prompt, the agent performed data auditing, literature-informed augmentation design, backbone and hyperparameter search, training, calibration, threshold selection, and final evaluation, without any manual coding by the investigators. All runs, hyperparameters, and metrics were tracked with an experiment-tracking dashboard, and every decision was recorded in a chronological log. Reproducibility was enforced using global seed 42, deterministic cuDNN, fixed CUBLAS workspace, seeded data loaders. Training was done using an NVIDIA A10G GPU, PyTorch 2.12.0, torchvision 0.27.0, and timm 1.0.27. The locked weights, preprocessing and calibration configurations, training and evaluation scripts and experiment log are openly archived in the model repository^16^.

#### Internal dataset and data audit

The internal dataset was TN5000,^14^an open-source thyroid nodule ultrasound dataset in which each nodule region of interest is cropped to a 224×224 RGB PNG. The predefined Train (n=3500), Valid (n=500), and Test (n=1000) splits were used unchanged, with label 1 denoting malignant nodules (the positive class). All three splits had a malignant majority (70.5%, 75.0%, and 73.1% malignant, respectively; Table 1). An automated audit found no corrupt images, exact pixel duplicates within or across splits, filename identifier overlap between splits, or label conflicts. Per-split mean pixel intensity was closely matched (∼81.4), indicating no detectable distribution shift or data leakage. The Train split was used for training only; the Valid split was used for model selection, calibration, and threshold selection. The Test split was used exactly once for final locked model evaluation.

**Table 1.** Characteristics of the internal (TN5000) and external (University of Colorado) datasets.

| **Characteristic** | **Internal: TN5000** | **External: University of Colorado** |
| --- | --- | --- |
| Unit of analysis | Nodule ROI crop | Nodule (paired transverse + longitudinal views) |
| Sample size | 5000 images (Train 3500 / Valid 500 / Test 1000) | 232 nodules |
| Malignant, n (%) | 3574 (71.5); 70.5 / 75 / 73.1 by split | 64 (27.6) |
| Benign, n (%) | 1426 (28.5) | 168 (72.4) |
| Reference standard | FNA cytology | Surgical histopathology or FNA cytology |
| Malignant histologies | Not subtyped | PTC 44, FV-PTC 12, MTC 5, FTC 2, HCC 1 |

*FNA, fine-needle aspiration; ROI, region of interest.*

#### Model development

ResNet-18 was used as the backbone^17^. The agent compared torchvision^18^and timm ImageNet-1k pretrained variants^19^ and used a single-logit classification head. A focused, 14-trial, one-factor at a time hyperparameter sweep was conducted around a literature-informed configuration, with the validation AUROC as the sole selection metric; the sweep varied the pretrained backbone, learning rate, weight decay, batch size, augmentation policy, class imbalance strategy, fine-tuning depth, and loss function. Augmentations were restricted to medically plausible B-mode ultrasound transforms (horizontal flip, rotation ≤10°, mild affine translation and scaling, ±15% brightness/contrast, light Gaussian blur), informed by ultrasound augmentation best practice^20^; vertical flip, large rotation or shear, aggressive cropping, and color jitter were deliberately avoided to preserve clinically relevant nodule morphology. Models were trained with the AdamW optimizer,^21^ a cosine-annealing schedule with warmup, early stopping on validation AUROC, and mixed precision.

### Calibration and threshold selection

After model selection, probability calibration was assessed on the validation set and corrected by temperature scaling.^22^The decision threshold was prespecified as the highest specificity threshold achieving a validation sensitivity of at least 0.95—a sensitivity-prioritized operating point chosen to approximate the expected sensitivity of FNA cytology. The threshold was locked before the internal test set was evaluated, and calibrated probabilities were used for all reported operating point metrics for the internal test set.

#### External validation dataset and processing

The locked model was externally validated on an independent cohort from the University of Colorado, originally assembled from genotyped participants in the Colorado Center for Personalized Medicine Biobank with thyroid nodules imaged using high-frequency^15^(10–17 MHz) ultrasound, a nodule diameter ≥10 mm, and a definitive diagnosis established by surgical histopathology or FNA cytology. We evaluated all nodules with paired transverse and longitudinal images and a definitive benign or malignant label, yielding 232 nodules. The locked preprocessing, temperature, and a test-time augmentation (TTA) scheme (original image plus horizontal flip plus eight mild random-affine passes, averaging calibrated probabilities) were applied unchanged. Because the validation derived locked threshold was developed on a different source, external operating point metrics were computed at the default probability cutoff of 0.50, while AUROC was used as the threshold-independent primary metric of discrimination.

#### Statistical analysis

Discrimination was summarized by AUROC, and classification performance evaluated by sensitivity, specificity, PPV, NPV; calibration was quantified by the expected calibration error (ECE) and Brier score. Ninety-five percent confidence intervals were estimated by stratified bootstrap resampling. For the external test set, performance was compared with a previously published BiT ResNet-50 classifier evaluated on the same University of Colorado source.^15^ The difference between models was assessed using the DeLong test.

## Results

### Datasets and autonomous workflow

Dataset characteristics for the internal and external cohorts are summarized in Table 1. The internal dataset comprised 5000 nodule-cropped images (3574 malignant, 1426 benign) across the predefined splits; the external cohort comprised 232 nodules (64 malignant) with diagnoses confirmed by histopathology or cytology and an enrichment of non-papillary histologies absent from most prior evaluations.

### Model selection and calibration

The best configuration achieved a validation AUROC of 0.98. The augmentation ablation confirmed the literature-informed default policy (validation AUROC 0.971–0.976) outperformed both a flip-only policy (0.964) and a stronger augmentation policy (0.961. The sweep was deliberately limited to 14 trials to avoid overfitting the 500-image validation set. Temperature scaling (T=0.565) improved validation calibration (ECE 0.083→0.031; Brier 0.059→0.052) while leaving AUROC unchanged. The prespecified rule produced a locked decision threshold of 0.711, at which validation sensitivity was 0.95 and specificity 0.9.

### Internal test performance

On the held out internal test set (n=1000), evaluated once with calibrated probabilities and the locked threshold, the model achieved an AUROC of 0.94 (95% CI, 0.920–0.953), sensitivity of 0.90, specificity of 0.8, PPV of 0.92, and NPV of 0.75 (Table 2). The model was also evaluated on the default threshold of 0.5 (Table 2).

**Table 2:** Internal test performance at two different thresholds.

| Metric | Threshold 0.71 (95% CI) | Threshold 0.5 |
| --- | --- | --- |
| Sensitivity | 0.90 (0.88–0.92) | 0.96 (0.94–0.97) |
| Specificity | 0.81(0.76–0.86) | 0.72 (0.66–0.77) |
| **PPV** | 0.93 (0.91–0.95) | 0.90 (0.89–0.92) |
| **NPV** | 0.75 (0.71–0.8) | 0.86 (0.82–0.90) |
| **AUROC** | 0.94 (0.93–0.96) | 0.94 (0.93–0.96) |

### External validation

On external per-nodule validation with TTA (the prespecified primary analysis), the model achieved an AUROC of 0.9 (95% CI, 0.85–0.94); at a 0.50 probability cutoff, sensitivity was 0.92, specificity 0.68, PPV 0.52, and NPV 0.96 (Table 3). Figure 1 depicts the confusion matrix for the agentic model’s performance on external test set.

**Figure 1:**
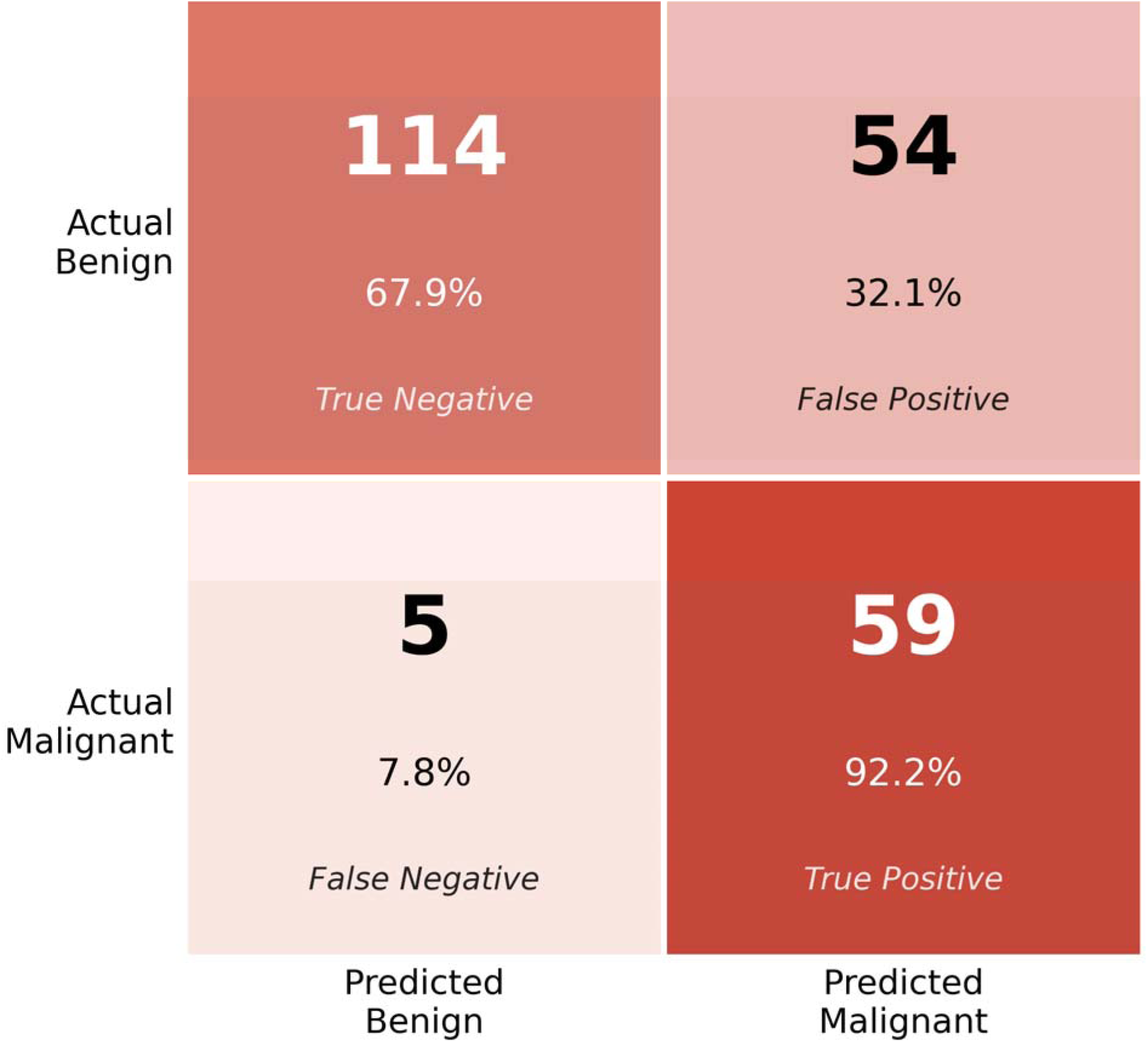
Confusion matrix for agentic model’s performance on external test set (N=232, threshold =0.5).

**Table 3:**
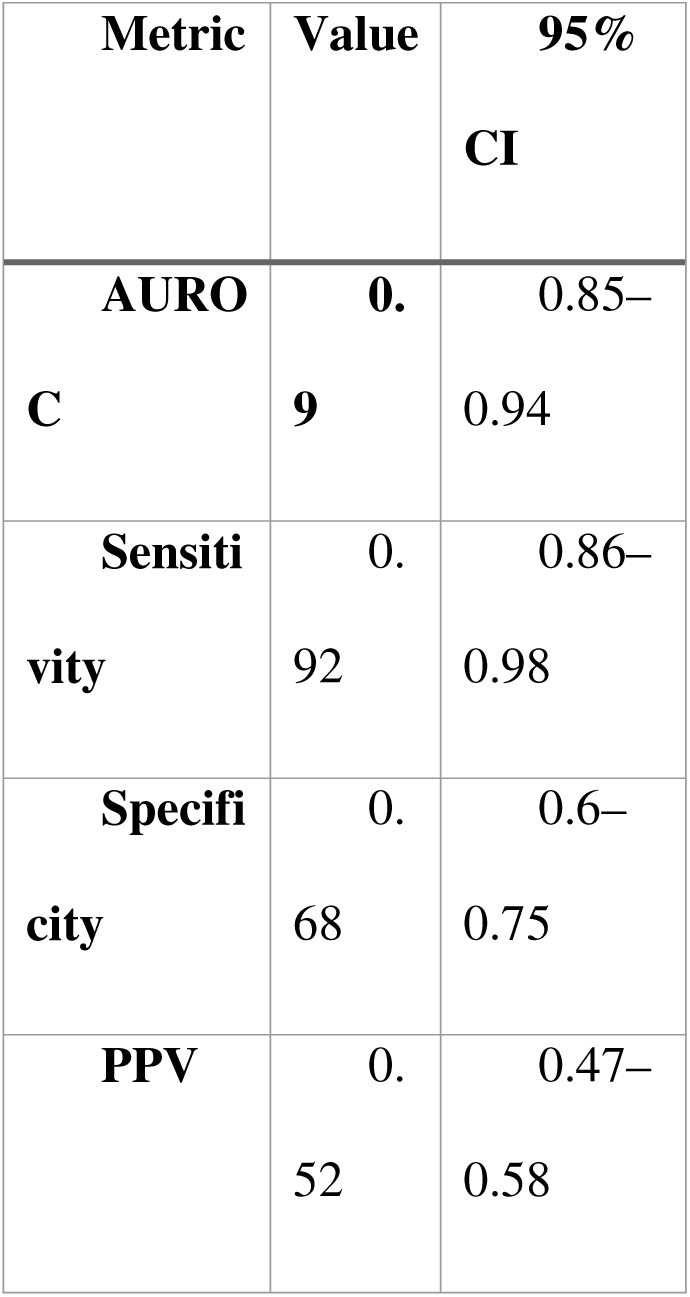
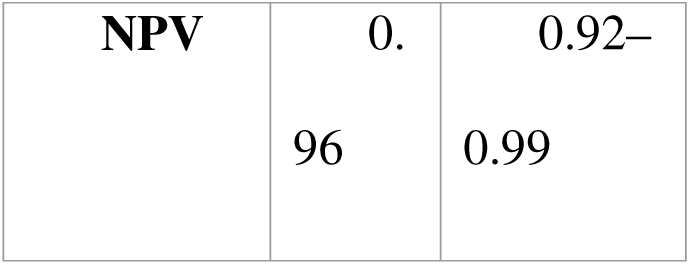
External validation results.

### Performance across thyroid cancer subtypes

Among the 64 malignant nodules, 59 (92%) were detected at the 0.50 cutoff. Detection rate was high for papillary carcinoma (43/44) oncocytic (1/1) and medullary (4/5) cancers, but lower for follicular pattern malignancies—FV-PTC (10/12) and FTC (1/2) (Figure 2). The missed cancers were concentrated among follicular-pattern tumors that characteristically lack the classic suspicious sonographic features exploited by image-based classifiers, consistent with prior observations.^15^

**Figure 2:**
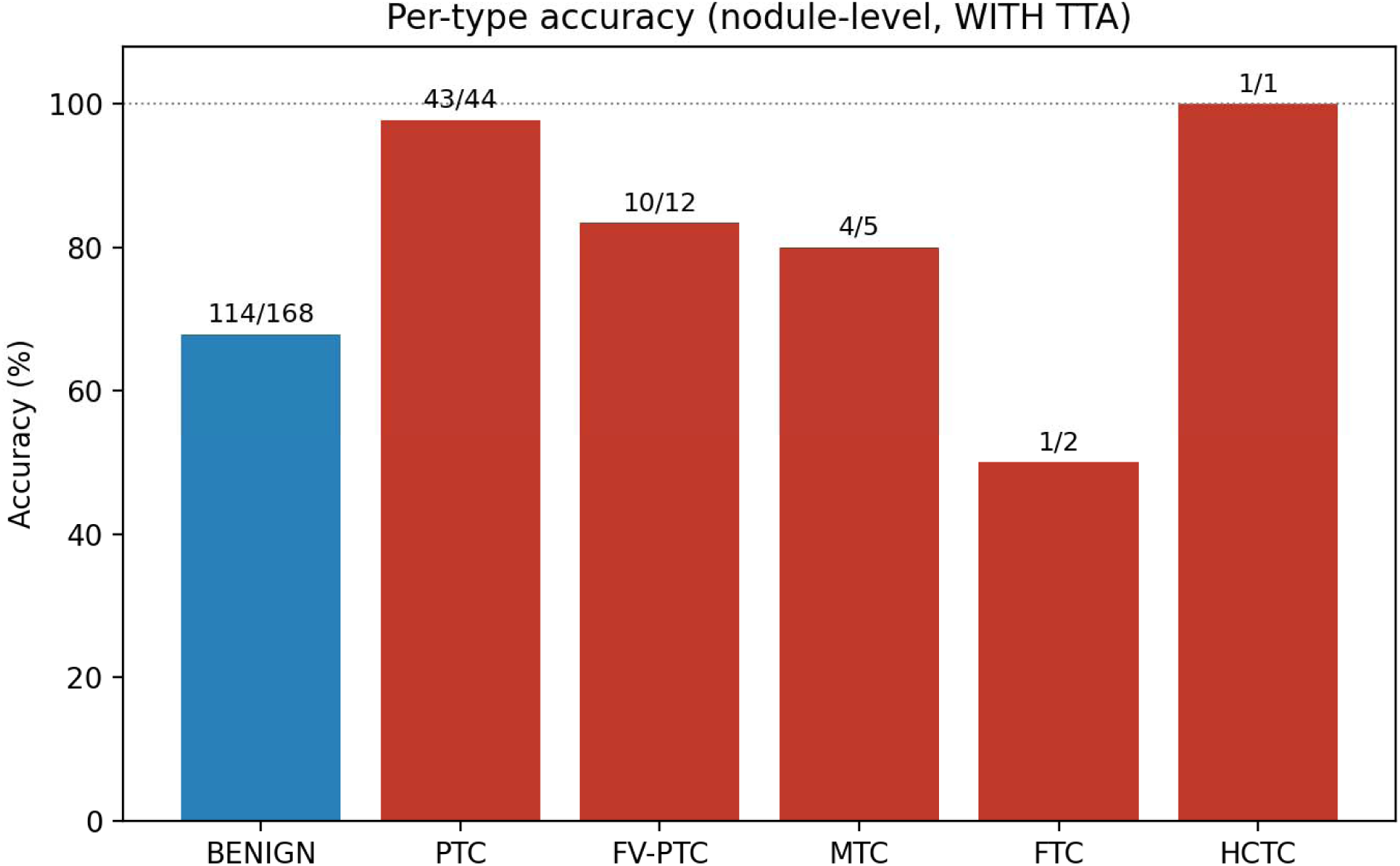
Performance of agentic model on different nodules subtypes in external test set.

### Benchmark comparison

On the same external source, a previously published BiT ResNet-50 classifier reported an AUROC of 0.83^15^. In the same dataset of 232 thyroid nodules (64 malignant [27.6%], 168 benign), the agentic-TTA model demonstrated better discrimination than the benchmark model with a smaller ResNet-18 model (Table 4). The area under the receiver-operating-characteristic curve (AUC) improved from 0.83 for the benchmark to 0.9 for the agentic TTA model (Figure 3). Paired DeLong test: 95% CI, +0.006 to +0.121; p = 0.029.

**Figure 3:**
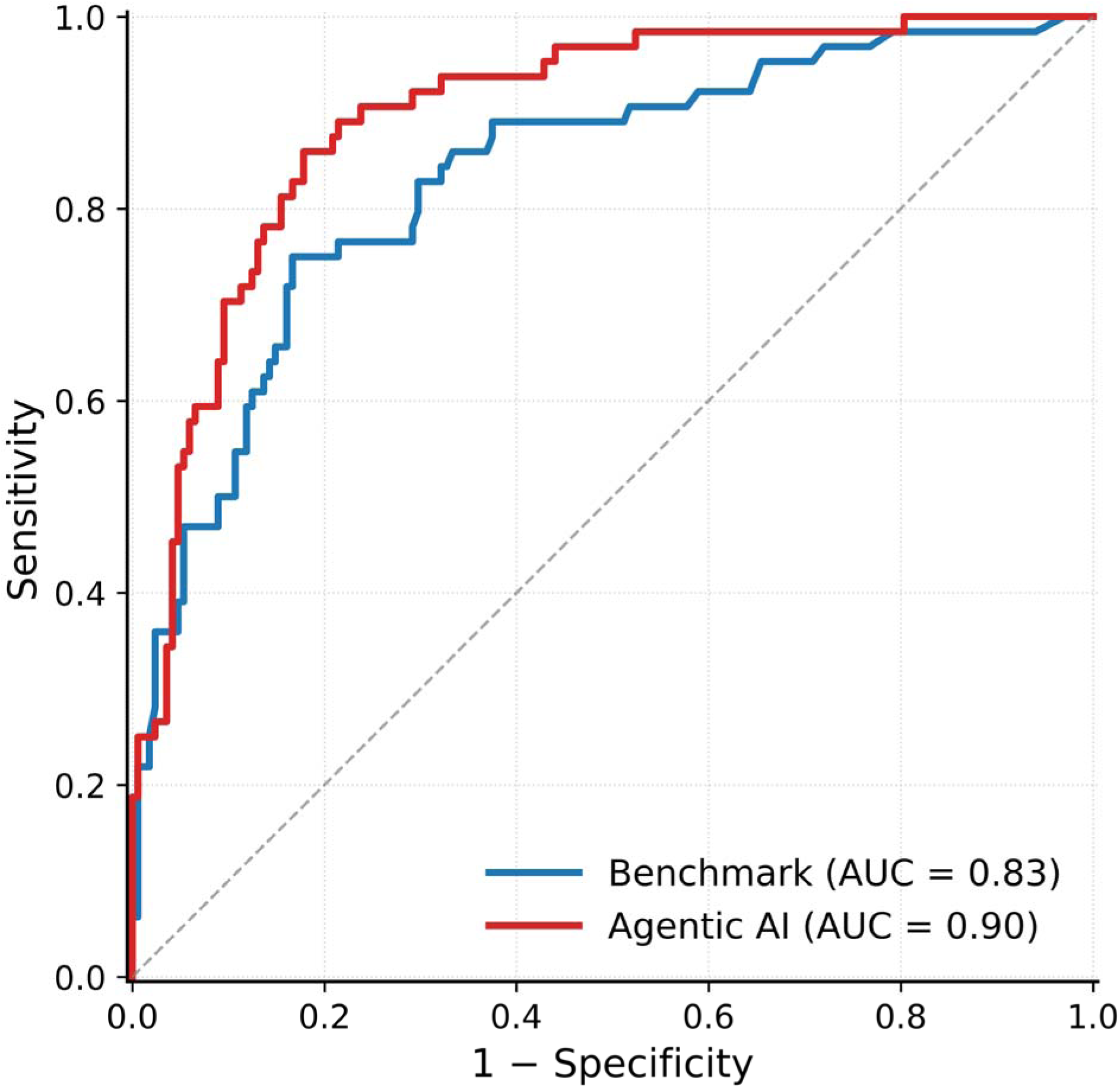
Receiver operating characteristic (ROC) curves for the benchmark model (blue) and the agentic AI model (red) on the external validation cohort of 232 thyroid nodules (64 malignant, 168 benign). The agentic AI model demonstrated higher discriminative performance (AUC = 0.90) than the benchmark model (AUC = 0.83). The diagonal dashed line represents the line of no discrimination. AUC, area under the curve.

**Table 4:**
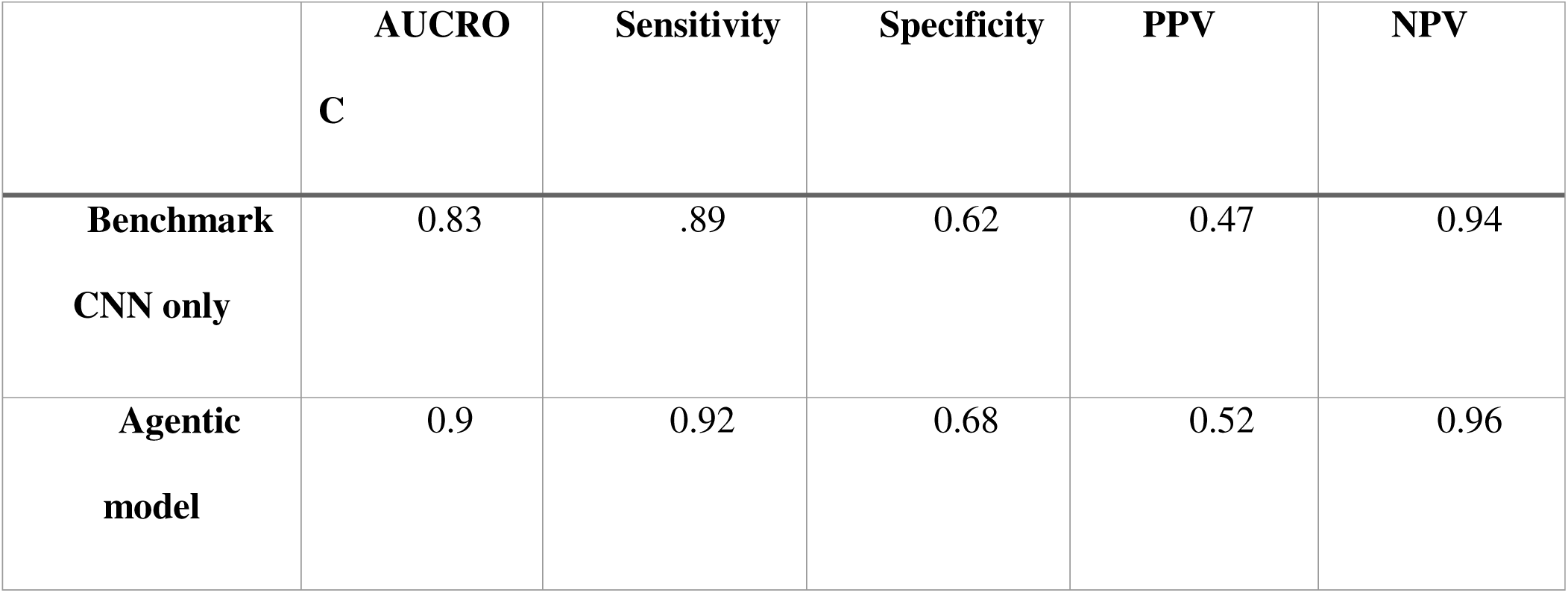
Comparison of performance of benchmark model against agentic model.

## Discussion

Using a publicly accessible, no-code agentic AI platform, we autonomously developed a calibrated thyroid nodule ultrasound classifier and subjected it to a prespecified, locked internal evaluation and an independent external validation. The model discriminated benign from malignant nodules well internally (AUROC 0.94) and generalized to an external cohort (AUROC 0.9) that included follicular, medullary, oncocytic, and follicular-variant of papillary thyroid cancers rarely represented in prior evaluations. To our knowledge, this is among the first demonstrations that an autonomous agent can produce a thyroid imaging classifier meeting the prespecified methodological expectations of a diagnostic study, data auditing, leakage checks, calibration, prespecified threshold locking, and external validation with confidence intervals.

The principal contribution is methodological; the entire pipeline was specified in a single natural language prompt and executed without manual coding. This directly addresses two barriers that have limited the clinical translation of thyroid AI: the scarcity of models that clinicians can independently test, and the machine-learning expertise required to create and run them. A no-code agentic route makes it feasible for clinician researchers who hold thyroid ultrasound data but do not write software to develop, calibrate, and validate their own models, and to do so reproducibly because every decision, configuration, and metric is logged. We are extending this work by deploying the locked model as a browser-based, on-device classifier (<u>ThyroNaut — Thyroid Nodule Ultrasound Classifier</u>) that runs locally without any external software installation or data upload, which would let endocrinologists test the model on their own images and enable the head-to-head comparisons the field currently lacks.

Sensitivity was high for papillary carcinoma and preserved for medullary and oncocytic tumors, but lower for follicular-pattern malignancies (FTC and FV-PTC), and the false-negative cases clustered among these subtypes. This mirrors the known limitation of image-based risk stratification, both by humans and machines – for cancers that lack classic suspicious features and underscores that orthogonal information (for example, molecular or genetic risk) may be needed to close this gap.^15^ Evaluating models on histologically diverse external data is essential, because performance measured only on papillary predominant cohorts overstates real-world generalizability.

In benchmarking on the same external source, the agentically developed ResNet-18 exceeded the performance range of a previously published BiT ResNet-50 classifier, despite its smaller size and image-only inputs^15^. This finding supports the feasibility of autonomous model development and the value of releasing a freely testable model as a common benchmark.

This study has important limitations. The model was trained on a single, cropped region of interest dataset; whole-frame ultrasound and images from other scanners or populations are out of distribution and would likely yield lower performance. The prespecified locked threshold was developed on the training source and was not transferred to the external cohort, where operating-point metrics were instead computed at a default 0.50 cutoff, and calibration drifted, so local recalibration is necessary for better performance. The external cohort was modest in size, and subtype subgroups were small. The evaluation was retrospective, not prospective; the workflow depends on a specific third-party agent platform. However, a similar workflow can be deployed on a local computer with enough computing power. There could be selection bias because only one transverse and longitudinal image of the nodule was used in the test set. We did not evaluate model performance on cine clips. Another limitation is that only nodules with FNA or histopathology results were included in the test set. This enriches suspicious lesions and inflates apparent discrimination relative to an unselected screening population. In the real world, all nodules seen in a clinical practice might be evaluated by this AI model, and performance might be different. We acknowledge that our model is proof-of-concept model and can be improved. We hope that the general research community uses our model as a baseline comparator and employs the reproducible framework and larger datasets to push the field forward.

In conclusion, an agentic, no-code AI workflow produced a calibrated, externally validated thyroid nodule ultrasound classifier that exceeded the performance of the benchmark model on the same external source despite using a lower parameter count. These results support a more accessible, reproducible, and independently testable model for medical AI development in endocrinology. Prospective, multisite validation, local recalibration will be required before clinical use.

## Data and Model Availability

The model code, locked weights, preprocessing, calibration configurations, experiment log are openly available in the Hugging Face model repository https://huggingface.co/Johnyquest7/agentic_thyroid_model. The TN5000 dataset is publicly available under a CC BY license. The external University of Colorado data are described in the original publication and is available at https://github.com/npozdey/thyroid_nodule_PRS. The model can be tested at <u>ThyroNaut — Thyroid Nodule Ultrasound Classifier</u>. Once the ResNet model downloads, you can disconnect the internet. It should still work without the internet. A YouTube video demonstrating the proper testing method is available at https://youtu.be/eUHhgsmiREY. The natural-language prompt that defined the agentic workflow is provided as Supplementary Material.

## Additional Information

### Author Contributions

JT conceived the study in discussion with NP. JT designed and supervised the agentic workflow, interpreted the results, and drafted the manuscript. NP contributed to data curation, external validation, and critical revision. All authors approved the final version.

## Data Availability

The model code, locked weights, preprocessing, calibration configurations, experiment log are openly available in the Hugging Face model repository https://huggingface.co/Johnyquest7/agentic_thyroid_model. The TN5000 dataset is publicly available under a CC BY license. The external University of Colorado data are described in the original publication and is available at https://github.com/npozdey/thyroid_nodule_PRS.

https://huggingface.co/datasets/Johnyquest7/TN5000-thyroid-nodule-classification

## Acknowledgments

The authors acknowledge the Hugging Face ML-Intern platform used for autonomous model development. We also acknowledge the creators of TN5000 datasets and open source python libraries that made this research possible.

## Abbreviations

ATA: American Thyroid Association
AUROC: area under the receiver-operating-characteristic curve
CNN: convolutional neural network
ECE: expected calibration error
FNA: fine-needle aspiration
FTC: follicular thyroid carcinoma
FV-PTC: follicular variant of papillary thyroid carcinoma
HCC: Hürthle (oncocytic) cell carcinoma
MTC: medullary thyroid carcinoma
NPV: negative predictive value
PPV: positive predictive value
PTC: papillary thyroid carcinoma
TI-RADS: Thyroid Imaging Reporting and Data System
TTA: test-time augmentation

## References

1. Peng Y, Wang TT, Wang JZ, et al. The application of artificial intelligence in thyroid nodules: a systematic review based on bibliometric analysis. Endocr Metab Immune Disord Drug Targets. 2024;24(11):1280–1290. doi:10.2174/0118715303264254231117113456

2. Sant VR, Radhachandran A, Ivezic V, et al. From bench-to-bedside: how artificial intelligence is changing thyroid nodule diagnostics, a systematic review. J Clin Endocrinol Metab. 2024;109(7):1684–1693. doi:10.1210/clinem/dgae277

3. Pozdeyev N, White SL, Bell CC, Haugen BR, Thomas J. Artificial intelligence applications in thyroid cancer care. J Clin Endocrinol Metab. 2026;111(2):316–324. doi:10.1210/clinem/dgaf530

4. Haugen BR, Alexander EK, Bible KC, et al. 2015 American Thyroid Association management guidelines for adult patients with thyroid nodules and differentiated thyroid cancer: the American Thyroid Association Guidelines Task Force on Thyroid Nodules and Differentiated Thyroid Cancer. Thyroid. 2016;26(1):1–133. doi:10.1089/thy.2015.0020

5. Tessler FN, Middleton WD, Grant EG, et al. ACR Thyroid Imaging, Reporting and Data System (TI-RADS): white paper of the ACR TI-RADS Committee. J Am Coll Radiol. 2017;14(5):587–595. doi:10.1016/j.jacr.2017.01.046

6. Hoang JK, Middleton WD, Farjat AE, et al. Interobserver variability of sonographic features used in the American College of Radiology Thyroid Imaging Reporting and Data System. AJR Am J Roentgenol. 2018;211(1):162–167. doi:10.2214/AJR.17.19192

7. Grussendorf M, Ruschenburg I, Brabant G. Malignancy rates in thyroid nodules: a long-term cohort study of 17,592 patients. Eur Thyroid J. 2022;11(4):e220027. doi:10.1530/ETJ-22-0027

8. Li X, Zhang S, Zhang Q, et al. Diagnosis of thyroid cancer using deep convolutional neural network models applied to sonographic images: a retrospective, multicohort, diagnostic study. Lancet Oncol. 2019;20(2):193–201. doi: 10.1016/S1470-2045(18)30762-9

9. Buda M, Wildman-Tobriner B, Hoang JK, et al. Management of thyroid nodules seen on US images: deep learning may match performance of radiologists. Radiology. 2019;292(3):695–701. doi: 10.1148/radiol.2019181343

10. Thomas J, Tessler FN. Artificial intelligence applications in thyroid cancer diagnosis: 2026 update. Thyroid. 2026;36(2):133–140. doi:10.1177/10507256251412316

11. Gatta E, Gatta R, Morandi R, et al. Machine learning for diagnosis of malignant thyroid nodules based on thyroid ultrasound: systematic review and meta-analysis of studies with external datasets. Eur J Radiol Open. 2025;16:100716. doi:10.1016/j.ejro.2025.100716

12. Trirat P, Jeong W, Hwang SJ. AutoML-Agent: a multi-agent LLM framework for full-pipeline AutoML. In: Proceedings of the 42nd International Conference on Machine Learning. Vol 267. PMLR; 2025:60099-60146.

13. Reedi AJ, Bonamy H, Di Cosmo Y, von Werra L, Tunstall L. ml-intern: an agent that autonomously researches, writes, and ships good quality ML related code using the Hugging Face ecosystem. Computer software. 2026.

14. Zhang H, Liu Q, Han X, et al. TN5000: an ultrasound image dataset for thyroid nodule detection and classification. Sci Data. 2025;12:1437. doi:10.1038/s41597-025-05757-4

15. Pozdeyev N, Dighe M, Barrio M, et al. Thyroid cancer polygenic risk score improves classification of thyroid nodules as benign or malignant. J Clin Endocrinol Metab. 2024;109(2):402–412. doi:10.1210/clinem/dgad530

16. Thomas J. agentic_thyroid_model. Computer software. Revision e4f94ea. Hugging Face; 2026. doi:10.57967/hf/9282

17. He K, Zhang X, Ren S, Sun J. Deep residual learning for image recognition. In: Proceedings of the IEEE Conference on Computer Vision and Pattern Recognition (CVPR). 2016:770–778. doi:10.1109/CVPR.2016.90

18. PyTorch Contributors. torchvision.models.resnet18. PyTorch documentation. Accessed June 20, 2026. https://docs.pytorch.org/vision/stable/models/generated/torchvision.models.resnet18.html

19. Wightman R. PyTorch Image Models (timm). GitHub; 2019. doi:10.5281/zenodo.4414861

20. Qi X, et al. MediAug: exploring visual augmentation in medical imaging. In: Ali S, Hogg DC, Peckham M, eds. Medical Image Understanding and Analysis. MIUA 2025. Lecture Notes in Computer Science. Vol 15916. Springer; 2026. doi:10.1007/978-3-031-98688-8_16

21. Loshchilov I, Hutter F. Decoupled weight decay regularization. In: Proceedings of the International Conference on Learning Representations (ICLR). 2019.

22. Guo C, Pleiss G, Sun Y, Weinberger KQ. On calibration of modern neural networks. In: Proceedings of the 34th International Conference on Machine Learning. PMLR; 2017;70:1321–1330.

