## Supplementary materials for "An Agentic, No Code Artificial Intelligence Workflow for Developing and Externally Validating a Thyroid Nodule Ultrasound Malignancy Classifier"

**Initial prompt**

This prompt was submitted to ChatGPT 5.5 to generate the final prompt to be entered into Huggingface ML-Intern space.

I have a Hugging Face dataset: <https://huggingface.co/datasets/Johnyquest7/TN5000-thyroid-nodule-classification/tree/main>

This contains thyroid ultrasound images with diagnoses. The columns are image and label, where label 1 means malignant and label 0 means benign. The main data folder at the above URL has three subfolders: Train, Valid, and Test.

I want to train a thyroid nodule classification model using this dataset. Please proceed as follows:

Data exploration

- First, analyze the dataset for class imbalance and generate a data exploration report.

Data splits

- Use the Train folder for training, the Valid folder for validation, and the Test folder for testing.

Model and training

- Use a ResNet-18 architecture.
- Search for the best image augmentation techniques and training parameters specifically for ultrasound imaging and, more specifically, thyroid nodule classification.
- Train the model and find the best-performing ResNet-18 configuration for this task.
- Optimize for the best AUC.
- Ensure the model is properly calibrated.

Thresholding and evaluation

- After training, use the validation dataset to find the best threshold for sensitivity optimization.
- Apply that threshold and evaluate the model on the test dataset.
- We also have an external dataset on which we plan to test the final model.

Metrics to report

- Sensitivity, specificity, NPV, PPV, and AUC-ROC.
- Provide 95% confidence intervals for the final metrics.
- Include a confusion matrix on the test dataset.
- All metrics should be publication-ready.

Experiment tracking

- Track the experiment using Trackio.
- Create a Trackio dashboard at Johnyquest7/Trackio_agentic_thyroid.

Reproducibility (critical)

- This experiment must be completely reproducible, as we plan to publish the results in a medical journal.
- Log all decisions into a log.md file in the model repo.
- Save the Python training script into the model repo.
- The goal is to reproduce this locally.

Model output

- Once training is complete, push the model to a new repo called Johnyquest7/agentic_thyroid_model.

I will load my Hugging Face account with enough credits to run this on GPU.

Now write a prompt for this to be inputted into Ml_Intern huggingface space.

**Verbatim development prompt passed into Hugging Face ML-Intern**

*The following natural-language prompt was provided to the ML-Intern agent and constitutes the prespecified analysis plan; it is reproduced verbatim.*

I want you to run a complete, reproducible medical imaging machine-learning experiment for thyroid nodule malignancy classification using my Hugging Face dataset:

Dataset: https://huggingface.co/datasets/Johnyquest7/TN5000-thyroid-nodule-classification/tree/main

Dataset repo ID: Johnyquest7/TN5000-thyroid-nodule-classification

Task: binary thyroid ultrasound image classification

Columns: image and label

Label meaning: 0 = benign, 1 = malignant

Folder/split structure: Train, Valid, Test

Use:

Train folder only for model training

Valid folder only for validation, hyperparameter/model-selection, calibration, and threshold selection

Test folder only once for final locked evaluation

Before proceeding with training, ask me any critical questions that would affect reproducibility, Hugging Face authentication, repo visibility, GPU hardware choice, or publication reporting. Do not ask questions that can be answered by inspecting the dataset or code. After questions are answered, proceed.

Main objective:

Train the best possible ResNet-18-based thyroid ultrasound malignancy classifier optimized primarily for validation AUROC, while also ensuring proper calibration and publication-ready reporting.

Important: This experiment is intended for a medical journal publication, so every decision must be logged and reproducible.

Required outputs:

A new Hugging Face model repo:

Johnyquest7/agentic_thyroid_model

A Trackio dashboard Space:

Johnyquest7/Trakio_agentic_thyroid

Use the official trackio Python package for experiment tracking. Log all runs, hyperparameters, augmentation settings, losses, AUROC, calibration metrics, thresholding results, and final test metrics.

In the final model repo, save:

train.py: full reproducible training script

evaluate.py: final evaluation script for validation/test splits

evaluate_external.py: script that can evaluate a future external dataset with the same preprocessing and the locked final threshold

requirements.txt

README.md model card

LOG.md: detailed chronological experiment log with all decisions, rationale, failed runs, selected run, calibration method, thresholding method, and final metrics

data_exploration_report.md

results/ folder containing all figures and tables

final trained model weights

final preprocessing configuration

final threshold configuration

calibration object/parameters if applicable

JSON/CSV files containing per-image predictions for validation and test sets

Dataset exploration requirements:

First inspect the dataset directly. Generate a publication-ready data_exploration_report.md with:

Number of images in Train, Valid, Test

Number and percentage benign/malignant in each split

Class imbalance ratio in each split

Image dimensions, channels, file format summary

Missing/corrupt image check

Duplicate image check within and across splits if feasible

Representative benign and malignant image grids from each split

Pixel intensity distribution summary

Any dataset leakage concerns

A clear statement confirming Train/Valid/Test were kept separate

Model requirements:

Use ResNet-18 as the backbone. Search and compare the best available ResNet-18 implementations/pretrained weights suitable for transfer learning. Include at minimum:

torchvision.models.resnet18(weights=ResNet18_Weights.DEFAULT)

timm ResNet-18 pretrained variants if available, such as resnet18.a1_in1k, resnet18.a2_in1k, or other current timm ResNet-18 ImageNet-pretrained variants

Select the final ResNet-18 version based on validation AUROC, not test performance.

Training requirements:

Use PyTorch. Make training fully deterministic as much as possible:

Set global seed

Log seed

Log package versions

Log hardware/GPU info

Log CUDA/cuDNN settings

Save all configs as YAML/JSON

Save exact command line used to run training

Save Git/Hugging Face commit hashes where possible

Use a clean, publication-ready training pipeline:

Binary classification output with one logit

Loss: compare BCEWithLogitsLoss with class weighting and optionally focal loss if class imbalance is significant

Optimizer search: AdamW preferred; compare with SGD only if time allows

Scheduler: cosine annealing or ReduceLROnPlateau

Early stopping based on validation AUROC

Mixed precision if GPU supports it

Save the best checkpoint by validation AUROC

Do not tune on the test set

Augmentation and preprocessing requirements:

Before finalizing augmentations, search recent literature and best practices for ultrasound image classification and thyroid nodule classification. Use medically plausible augmentations only. Evaluate a small ablation of augmentation policies.

Candidate augmentations to consider:

Resize/crop to 224×224 if needed

Horizontal flip if anatomically acceptable

Small rotations, e.g., ±10–15 degrees

Small translations/scale/affine transforms

Brightness/contrast/gamma adjustment

Mild Gaussian noise or speckle-like noise

Random resized crop with conservative scale limits

CLAHE or histogram equalization only as an ablation, not blindly

Avoid augmentations that distort clinically relevant nodule morphology or ultrasound texture too aggressively

For validation/test, use deterministic preprocessing only. No augmentation.

Hyperparameter search:

Run a focused search optimized for validation AUROC. Search should include:

ResNet-18 pretrained variant

Learning rate

Weight decay

Batch size

Augmentation policy

Class imbalance strategy

Number of frozen vs unfrozen layers, or full fine-tuning

Scheduler choice if feasible

Use a reasonable GPU-aware search strategy. Do not overfit the validation set with excessive trial counts. Log all trials to Trackio.

Calibration requirements:

After selecting the best model by validation AUROC, assess calibration on the validation set:

Reliability diagram

Expected calibration error, ECE

Brier score

Calibration curve

Compare uncalibrated probabilities vs calibrated probabilities

Use temperature scaling or Platt scaling on the validation set if it improves calibration without harming discrimination

Save calibration parameters

Use the calibrated probabilities for threshold selection and final test reporting unless calibration clearly fails

Threshold selection:

After model selection and calibration, use the validation set to choose a locked decision threshold optimized for sensitivity. The thresholding method should be explicitly documented.

Use the following approach unless there is a strong reason not to:

Choose the highest-specificity threshold that achieves a target sensitivity of at least 0.95 on the validation set.

If 0.95 sensitivity is not achievable or unstable, choose the threshold that maximizes sensitivity while maintaining clinically reasonable specificity, and document the reason.

Also report Youden-index threshold as a secondary/reference threshold, but do not use it as the main threshold unless sensitivity optimization fails.

Lock the chosen threshold before evaluating the test set.

Final test evaluation:

Evaluate the locked final model once on the Test split using:

Calibrated probabilities

Locked validation-derived threshold

Report:

Sensitivity

Specificity

PPV

NPV

AUROC

Accuracy

F1 score

Brier score

ECE

Confusion matrix

ROC curve

Precision-recall curve

Calibration plot

Per-image prediction CSV with image identifier, true label, probability, predicted label

Publication-ready 95% confidence intervals:

For final Test metrics, calculate 95% CIs:

AUROC: bootstrap CI, preferably stratified bootstrap with at least 2000 resamples

Sensitivity/specificity/PPV/NPV/accuracy/F1: bootstrap CI or exact/binomial CI where appropriate

Clearly document CI method

Set and log bootstrap seed

Include point estimates and 95% CI in a clean markdown and CSV table

Confusion matrix:

Generate and save:

Raw count confusion matrix

Normalized confusion matrix

Publication-ready figure with labels:

True benign

True malignant

Predicted benign

Predicted malignant

External dataset readiness:

We have an external dataset that we plan to evaluate later. Create evaluate_external.py so that I can later run:

python evaluate_external.py \

--model_repo Johnyquest7/agentic_thyroid_model \

--data_dir /path/to/external_dataset \

--output_dir external_results

The external dataset script should support either:

folder format with benign/malignant subfolders, or

CSV with image paths and labels

It must use the same preprocessing, calibration, and locked threshold from the final model repo.

Reproducibility requirements:

Create a single-command local reproduction workflow. The repo should allow me to reproduce training locally with:

pip install -r requirements.txt

python train.py --config configs/final_config.yaml

python evaluate.py --split test --config configs/final_config.yaml

Also provide a command for re-running data exploration:

python explore_data.py --dataset_id Johnyquest7/TN5000-thyroid-nodule-classification

LOG.md requirements:

The LOG.md file must include:

Date/time of experiment

Dataset source and version/commit if available

Exact split usage

Class distribution

Literature-informed augmentation rationale

All model variants tried

All hyperparameters tried

Validation AUROC for each run

Calibration decision

Threshold selection decision

Final locked threshold

Final test results with 95% CIs

Limitations

Notes about external validation not yet performed

Statement that test set was evaluated only after model/threshold/calibration were locked

Model card requirements:

The README.md model card must include:

Intended use: research only, not clinical deployment

Dataset description

Label definitions

Preprocessing

Model architecture

Training procedure

Validation threshold strategy

Test performance table with 95% CIs

Calibration results

Confusion matrix image

Limitations and bias/leakage concerns

Warning that this model requires external validation before clinical use

Trackio requirements:

Use trackio for experiment tracking:

Project name: agentic_thyroid_resnet18

Space/dashboard: Johnyquest7/Trakio_agentic_thyroid

Log train/validation loss, AUROC, sensitivity, specificity, PPV, NPV, ECE, Brier score, learning rate, epoch, and hyperparameters

Log artifacts/figures if supported

Make sure final run is clearly marked as the selected model

Deliverables summary:

At the end, provide:

Link to the model repo: Johnyquest7/agentic_thyroid_model

Link to the Trackio dashboard: Johnyquest7/Trakio_agentic_thyroid

Final selected ResNet-18 variant

Final augmentation policy

Final training parameters

Final calibration method

Final validation-derived threshold

Final Test metrics with 95% CIs

Confusion matrix

Instructions to reproduce locally

Any limitations or failed checks

Do not push incomplete or untested code. If training cannot be completed because of missing permissions, insufficient compute, API/token issues, or package problems, stop and report exactly what failed, what was completed, and what command I should run next.
